# Mental Health of International Migrant Workers Amidst Large-Scale Dormitory Outbreaks of COVID-19: A Population Survey

**DOI:** 10.1101/2021.02.24.21252414

**Authors:** Young Ern Saw, Edina YQ Tan, P Buvanaswari, Kinjal Doshi, Jean CJ Liu

## Abstract

**Background:** In the COVID-19 pandemic, international migrant workers have faced increased vulnerability on account of their status. This study examined the mental health burden of COVID-19 amongst low-waged migrant workers involved in large-scale dormitory outbreaks within Singapore.

**Methods:** Between 22 June to 11 October 2020, questionnaires were distributed in-person and online to 1011 migrant workers undergoing movement restrictions. Mental health symptoms were measured using the 21-item Depression, Anxiety and Stress Scale (DASS-21). As covariates, we assessed participants’ socio-demographics, quarantine status, COVID-19 health concerns, financial stability, and exposure to news and misinformation. Linear regression models were fitted to identify factors associated with each DASS-21 subscale.

**Findings:** Complete movement restrictions were associated with increased depression and stress symptoms, while being diagnosed with COVID-19 was associated with increased anxiety. Participants who harboured fears about their health or job, perceived their health to be poorer, or had greater exposure to COVID-19 rumours reported higher depression, anxiety, and stress levels. Across the cohort, rates of severe or extremely severe depression (3.1%, 95% CI: 2.1-4.3%), anxiety (4.1%, 95% CI: 2.9-5.5%), and stress (1.3%, 95% CI: 0.7-2.2%) were similar to those observed in the general population for the host country (Singapore).

**Interpretation:** The risk factors identified underscore how the ongoing pandemic may impact the mental health of migrant workers. At the same time, we observed resilience within the cohort, with no evidence of increased symptomology (relative to the general population).

**Funding:** JY Pillay Global Asia Grant

**Research in Context:** *Evidence before this study:* We searched PubMed and Google Scholar for articles published in English between Jan 1, 2020 and Feb 20, 2021 using the following keywords: (“COVID*” OR “coronavirus”) AND (“mental*” OR “psychiatr*”) AND (“labo*r migra*” OR “migrant work” OR “foreign-work” OR “immigrant work” OR “economic migra*” OR “economic immigra*”). Focusing on international migrant workers employed in low-wage manual labour positions, we identified commentaries and interview-based studies describing the stressors faced by workers during the COVID-19 pandemic. However, we found no study documenting mental health symptoms within this group.

*Added value of this study:* To our knowledge, this is the first mental health survey of low-wage migrant workers during the COVID-19 pandemic. We observed that the mental health burden was highest amongst participants who encountered pandemic-related adversities (complete movement restrictions, testing positive for COVID-19), perceived the situation negatively (being fearful of their health or job, or judging their health to be poorer), or had higher exposure to COVID-19 rumours.

*Implications of the available evidence:* Our findings provide a basis to identify and support at-risk migrant workers during the pandemic. Although we did not observe elevated rates of depression, anxiety, and stress symptoms within the migrant worker cohort, individual workers who experience poor mental health may find it harder to access health-care services (relative to the general population). Correspondingly, targeted support for at-risk migrant workers may serve to reduce mental health inequalities.

## Introduction

In this era of globalisation, 164 million people worldwide are international migrant workers employed outside their countries of birth.^1^ They constitute a fifth of the workforce in high-income countries, and more than 40% of the workforce in several territories (e.g., Gulf States).^1^ Nonetheless, migrant workers often have disproportionately less access to the healthcare, information, and resources of their host countries.^2,3^

In the current COVID-19 pandemic, migrant workers have faced increased vulnerability on account of their status.^4^ For workers in low-waged, manual labour jobs, COVID-19 transmission has been elevated – largely owing to the high-density work conditions (e.g., factories) and housing arrangements (e.g., dormitories, shared housing) that many workers operate in.^5,6^ Correspondingly, COVID-19 outbreaks have occurred amidst migrant worker communities in Singapore, Thailand, Malaysia, and the Gulf states,^6–8^ with Singapore reporting a disease prevalence rate 188 times higher amongst migrant workers (47%) than in the general community (0.25%).^9^

Despite this increased vulnerability, there has been no study documenting migrant workers’ mental health during the pandemic. Within the general population, several meta-analyses have identified quarantine status, COVID-19 health concerns, financial instability, exposure to news and misinformation, and demographics (gender, age, education) as risk factors for poor mental health.^10–12^ These factors are likely accentuated amongst migrant workers. For example, having borne the brunt of infection clusters, many migrant groups have come under extended quarantine protocols as governments acted to control the COVID-19 spread.^13–15^ Relative to the general population, migrant workers have also experienced heightened threats of financial instability, and decreased access to official COVID-19 information.^4–6^ Together, these factors emphasize the need for cohort studies assessing the mental health burden of COVID-19 within this population. Such studies are needed to guide government and employer policies, and to identify workers in greatest need of support amidst the ongoing pandemic.

In this study, we characterized the depression, anxiety, and stress symptoms of migrant workers in Singapore. The city-state has a workforce of nearly 400,000 male workers employed in low-waged, manual-labour positions within the construction, shipping, and process sectors.^8,14^ Workers come primarily from South Asia, and reside in dormitories that host up to 25,000 residents (averaging 10 persons per room).^8,14^ Given this high density, dormitories were the epicentre of COVID-19 transmission between April to August 2020, contributing more than 90% of Singapore’s 59,000 COVID-19 cases to date.^9^ Consequently, all dormitory residents have been placed under prolonged movement restrictions: commencing in April 2020 with in-room or in-dormitory quarantines, to the gradual resumption of work activities five months later (August 2020), to getting limited access to sites outside the dormitory on non-working days (current status, as of February 2021) (Figure 1).^9^ Under these circumstances, we evaluated the mental health impact of COVID-19. We addressed two primary aims: (1) to document mental health symptoms amongst migrant workers, and (2) to identify risk and protective factors amidst the pandemic.

**Figure 1.**
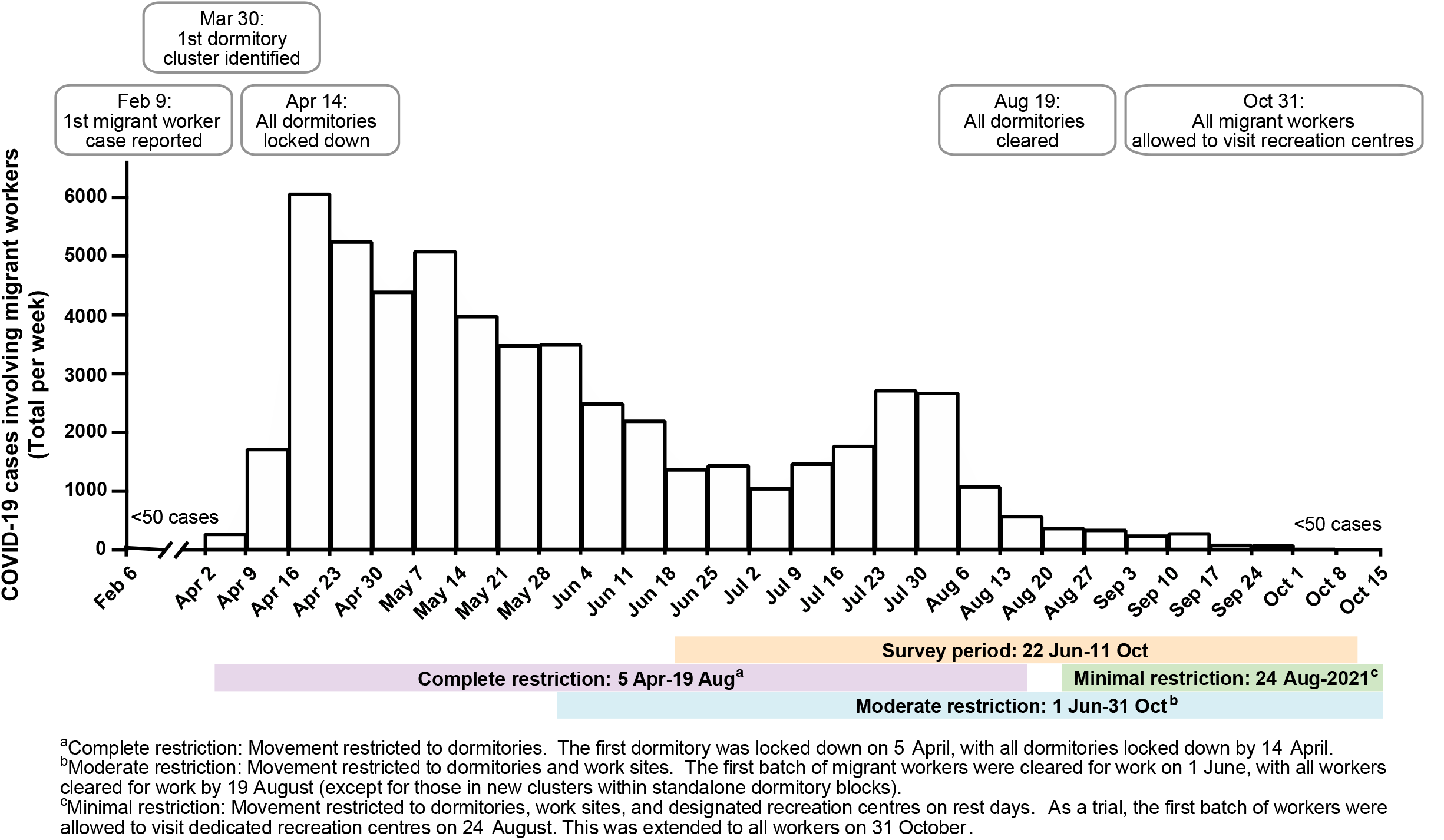
Chronology of COVID-19 events and case numbers within the migrant worker community in 2020.

## Methods

### Study design and participants

We used data from the COVID-19 Migrant Health Study, a cross-sectional survey of 1011 male migrants employed in manual labour jobs within Singapore. Participant eligibility criteria included: age ≥ 21 years, and holding a government work permit identifying their employment status. Data collection took place between 22 June to 11 October 2020 through two means. First, we conducted in-person surveys at: (1) a dormitory associated with Singapore’s largest COVID-19 cluster, (2) temporary accommodation for workers relocated from their dormitories, and (3) a recreation centre for workers. Second, we used physical posters and site-based messaging groups (on WhatsApp and Telegram) to advertise an online survey link in: (1) government quarantine facilities set up for active COVID-19 cases,^16^ and (2) dormitories housing migrant workers.^9,15^ Surveys were administered in participants’ native languages (English, Bengali, Tamil, or Mandarin), and were available in both written text and audio recordings to ensure access regardless of literacy level. Members of the trained research team were multi-lingual and multi-cultural.

All participants provided informed consent. Those who were recruited in person were reimbursed SGD $10. The study was approved by the Institutional Review Boards of the National University of Singapore and Singapore Health Services, and was pre-registered on ClinicalTrials.gov (NCT04448704).

#### Outcome measures

To assess psychological distress, we administered the 21-item Depression, Anxiety and Stress Scale (DASS-21).^17^ The DASS-21 is widely-used to screen for depression, anxiety, and stress symptoms, and has been used during the COVID-19 pandemic to capture mental health across countries and population segments (e.g., patients groups, healthcare workers, general population).^10,18^

Using the DASS-21, participants indicate the presence of symptoms over the past week, with each item rated on a 4-point scale ranging from “Never” (0) to “Almost Always” (3). Following convention, items were summed and multiplied by two to create a score for depression (7 items, range 0-42), anxiety (7 items, range 0-42), and stress (7 items, range 0-42). On the depression subscale, scores of 0-9 corresponded to normal levels, 10-13 to mild depression, 14-20 to moderate depression, 21-27 to severe depression, and ≥28 to extremely severe depression. For anxiety, 0-7 represented normal levels, 8-9 mild anxiety, 10-14 moderate anxiety, 15-19 severe anxiety, and ≥20 extremely severe anxiety. Finally, for the stress subscale, a score of 0-14 was in the normal range, 15-18 corresponded to mild stress, 19-25 to moderate stress, 26-33 to severe stress, and ≥34 to extremely severe stress.^17^

#### Covariates

As covariates, we included the following sociodemographic variables: age group (21-30, 31-40, 41-60), marital status (married, not married: single/widowed/separated/divorced), education (primary, secondary, tertiary), country of origin (Bangladesh, India, others), and years spent in Singapore (≤ 5 years, > 5 years). We also measured variables identified in previous studies as risk factors for poor mental health during the COVID-19 pandemic: namely, (i) quarantine status, (ii) COVID-19 health concerns, (iii) financial instability, and (iv) exposure to news and misinformation.^10–12^

Quarantine status was derived by matching government-imposed movement restrictions to the survey time-stamp and site (since restrictions differed across sites). Using this information, status was categorized as: ‘complete restriction’ (unable to leave the dormitory for any reason), ‘moderate restriction’ (movements restricted to the dormitory and to work sites), or ‘minimal restriction’ (movements restricted to the dormitory, work sites, and assigned recreation centres on rest days). Participants also self-reported their trust in the institutional response (‘confident that the government can control the spread of COVID-19’, ‘not confident that the government can control the spread of COVID-19’).

To capture COVID-19 health concerns, we documented participants’ history of COVID-19 (tested positive: yes, no), their self-rated health (poor / normal, good), and whether they were fearful about their health during the COVID-19 outbreak (afraid, not afraid).

Financial stability was captured by asking participants whether they were fearful about losing their job during the COVID-19 pandemic (afraid, not afraid), what their income was before and during the COVID-19 pandemic (current income categorized as: ‘less than before’, or ‘same / more than before’ the pandemic), whether they had debts to repay (yes / no), the number of family members dependent on their income (<5 family members, ≥5 family members), and whether they had a bank account in Singapore (yes / no).

Finally, to measure exposure to news and misinformation, participants indicated their familiarity with 5 common COVID-19 rumours (drinking water frequently will help to prevent infection; eating garlic can help to prevent infection; the outbreak came about because of people eating bat soup; the virus was created in America to affect China; and the virus was created in China as a weapon).^19^ The number of rumours participants recognized were summed and categorized as ‘low exposure’ (0-2 rumours) or ‘high exposure’ (3-5 rumours). Additionally, participants reported the amount of time they spent checking COVID-19 news (≤2 hours / day, *>*2 hours / day), and using social media to discuss or share information about COVID-19 (≤2 hours / day, *>*2 hours / day).

#### Statistical analysis

We first summarized participants’ baseline characteristics with counts (%) and medians (with interquartile ranges; IQR). For the primary analyses, we conducted linear regression models with each DASS-21 subscale score (depression, anxiety, and stress) as outcome measures. To identify risk factors for poor mental health, we included the full set of covariates as predictor variables (namely, socio-demographics, quarantine status, COVID-19 health concerns, financial stability, and exposure to news and misinformation).

For each regression model, the type 1 family-wise error rate was controlled at 0.05 through Bonferroni correction (Bonferroni-adjusted alpha level of 0.05 / 22 predictors = 0.002). All analyses were done using R (Version 4.0.3) and SPSS (Version 23.0).

#### Role of the funding source

The funders had no role in the design and conduct of the study; in data collection, analysis, and interpretation; or in manuscript preparation. All authors had full access to the data, and were responsible for the decision to submit the manuscript for publication.

## Results

### Study participants

Between 22 June to 11 October 2020, we surveyed 1011 migrant workers employed in manual labour positions within Singapore (78.5% response rate; Figure 2). As shown in Table 1, all participants were men with a median age of 32 years (IQR: 28-37). The large majority were from South Asia including Bangladesh (57.1%) and India (37.8%), had spent a median of 7 years in Singapore (IQR: 4-10), and worked in the construction sector (86.9%).

**Table 1.**
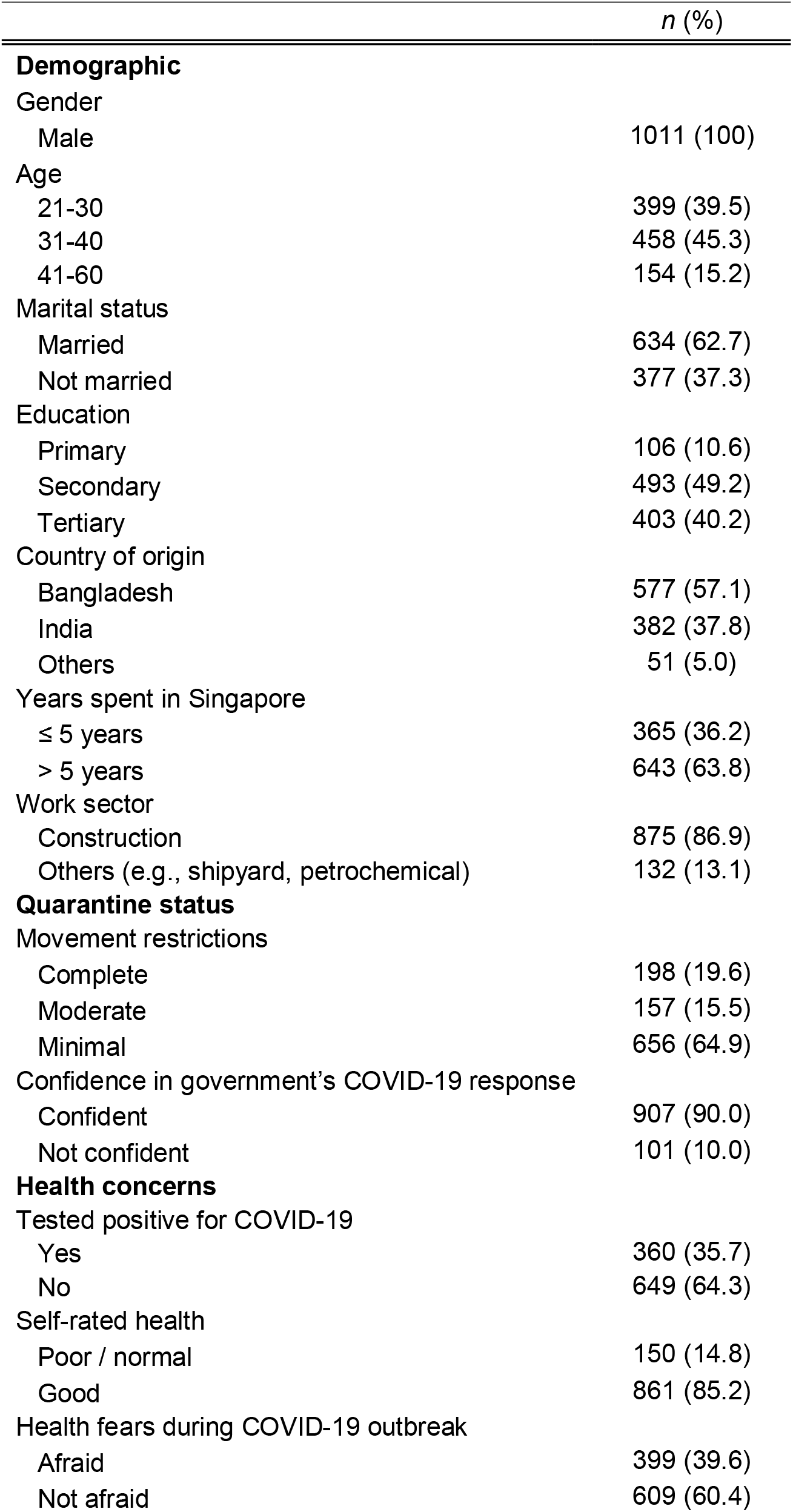
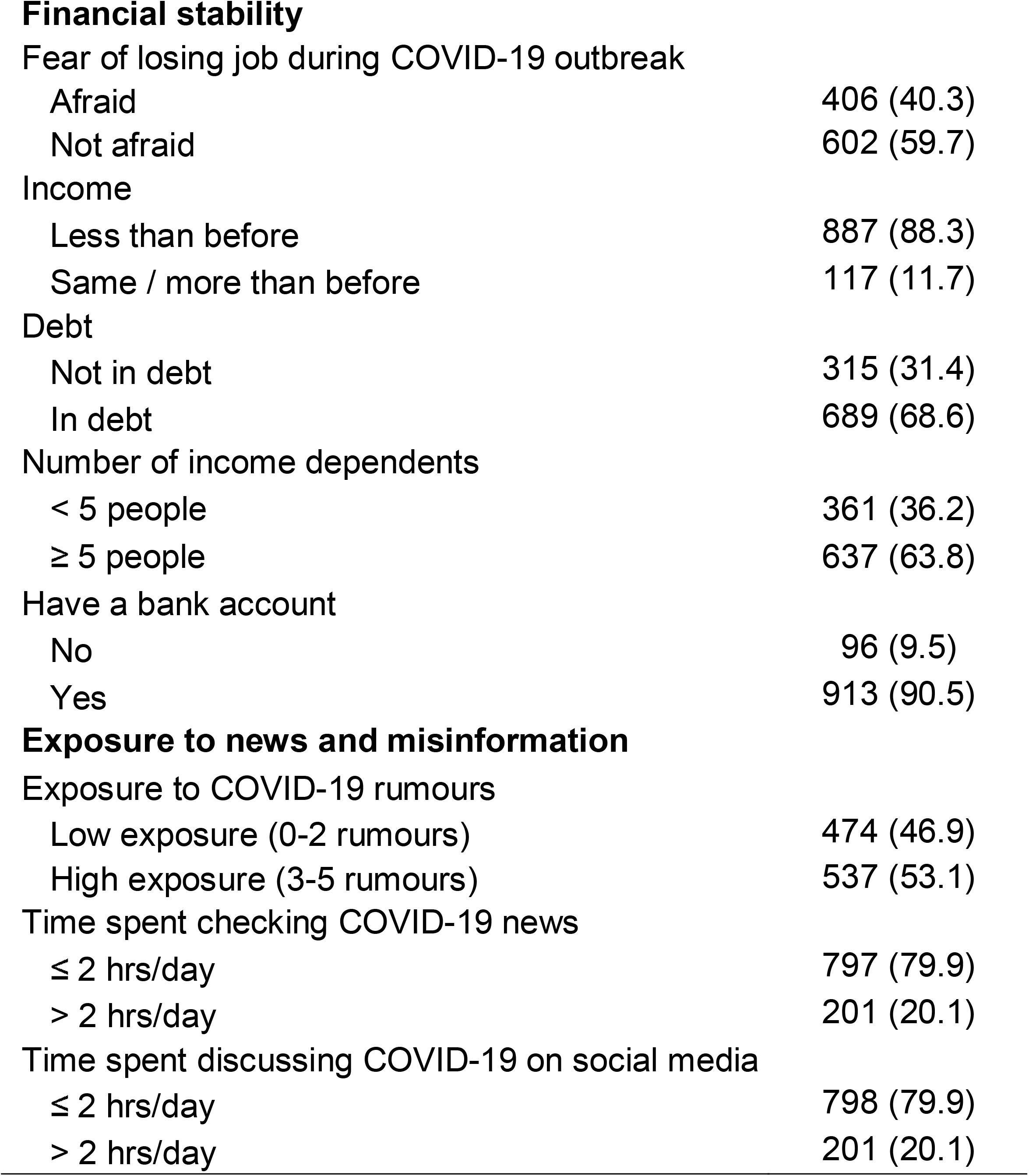
Profile of survey respondents (N = 1011)

**Figure 2.**
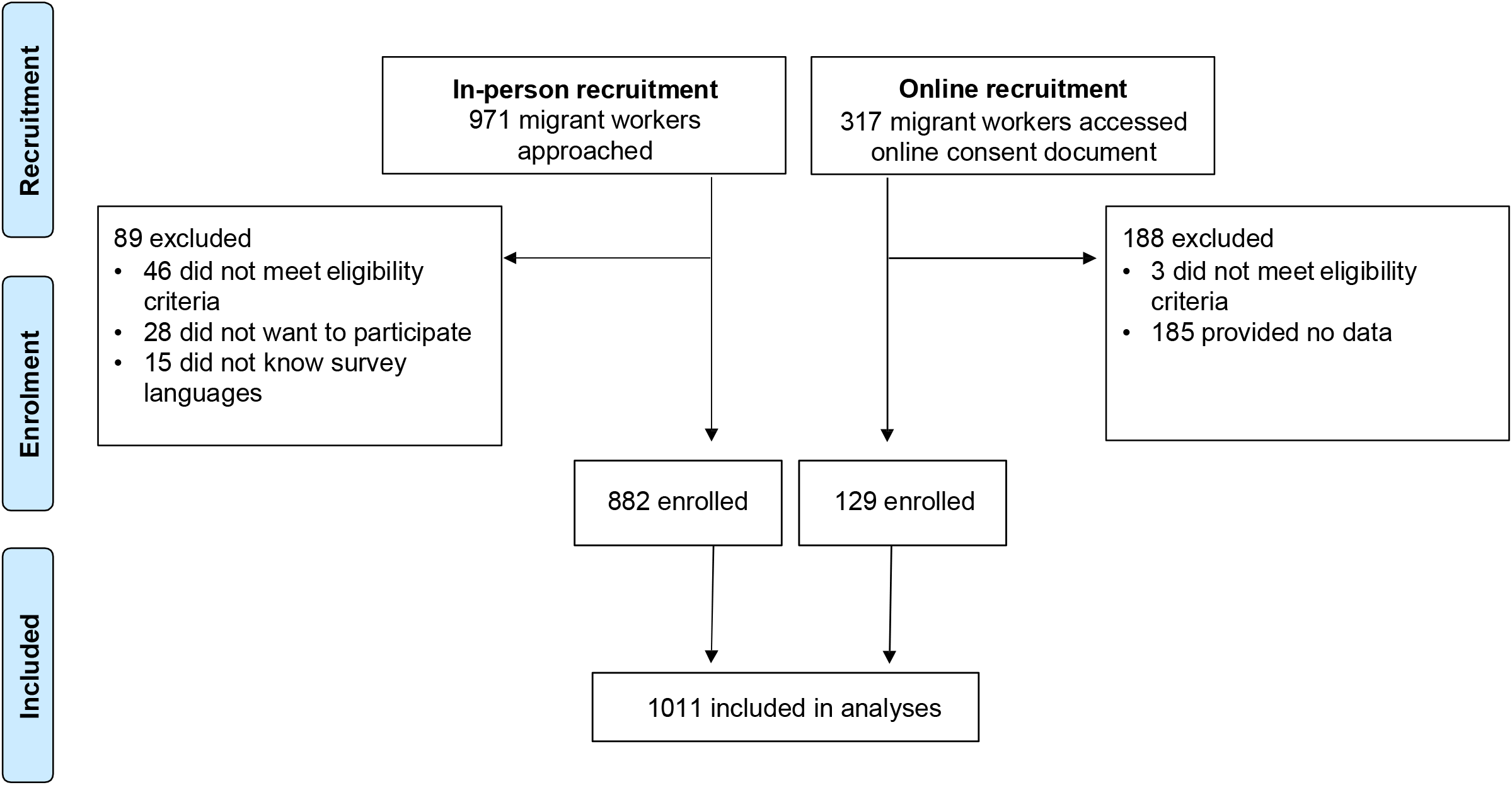
Flowchart of study methodology and participant inclusion.

#### Mental health symptoms of migrant workers

For the DASS-21 depression subscale, the median score was 4 (IQR: 0-8). 3.1% of participants (95% CI: 2.1-4.3%) met severe or extremely severe cut-offs, 8.6% (95% CI: 7.0-10.5%) met moderate cut-offs, and 9.6% (95% CI: 7.9-11.6%) met mild cut-offs. On the anxiety subscale, the median score was 2 (IQR: 0-4), with 4.1% (95% CI: 2.9-5.5%) reporting severe or extremely severe levels, 8.0% (95% CI: 6.4-9.9%) reporting moderate levels, and 5.4% (95% CI 4.1-7.0) reporting mild levels. Finally, on the stress subscale, participants reported a median score of 4 (IQR: 0-8) with 1.3% of participants (95% CI: 0.7-2.2%) experiencing severe or extremely severe stress, 1.9% (95% CI: 1.1-2.9%) experiencing moderate stress, and 3.4% (95% CI: 2.3-4.7%) experiencing mild stress. These rates are similar to those observed in community samples within Singapore during the COVID-19 pandemic (e.g., severe or extremely severe depression: 3.1% in our sample vs 5.3% in a population survey; anxiety: 4.1% vs 4.5%; stress: 1.3% vs 2.5%).^19^

#### Factors associated with mental health symptoms among migrant workers

Movement restrictions predicted depression and stress symptoms (Table 2 & Figure 3). Namely, higher subscale scores were observed amongst participants surveyed during complete movement restrictions, as compared to participants surveyed during minimal restrictions (depression: b = 2.38, p < 0.001; stress: b = 1.98, p < 0.001). We found no evidence that symptoms depended on participants’ self-reported trust in the institutional response.

**Table 2.**
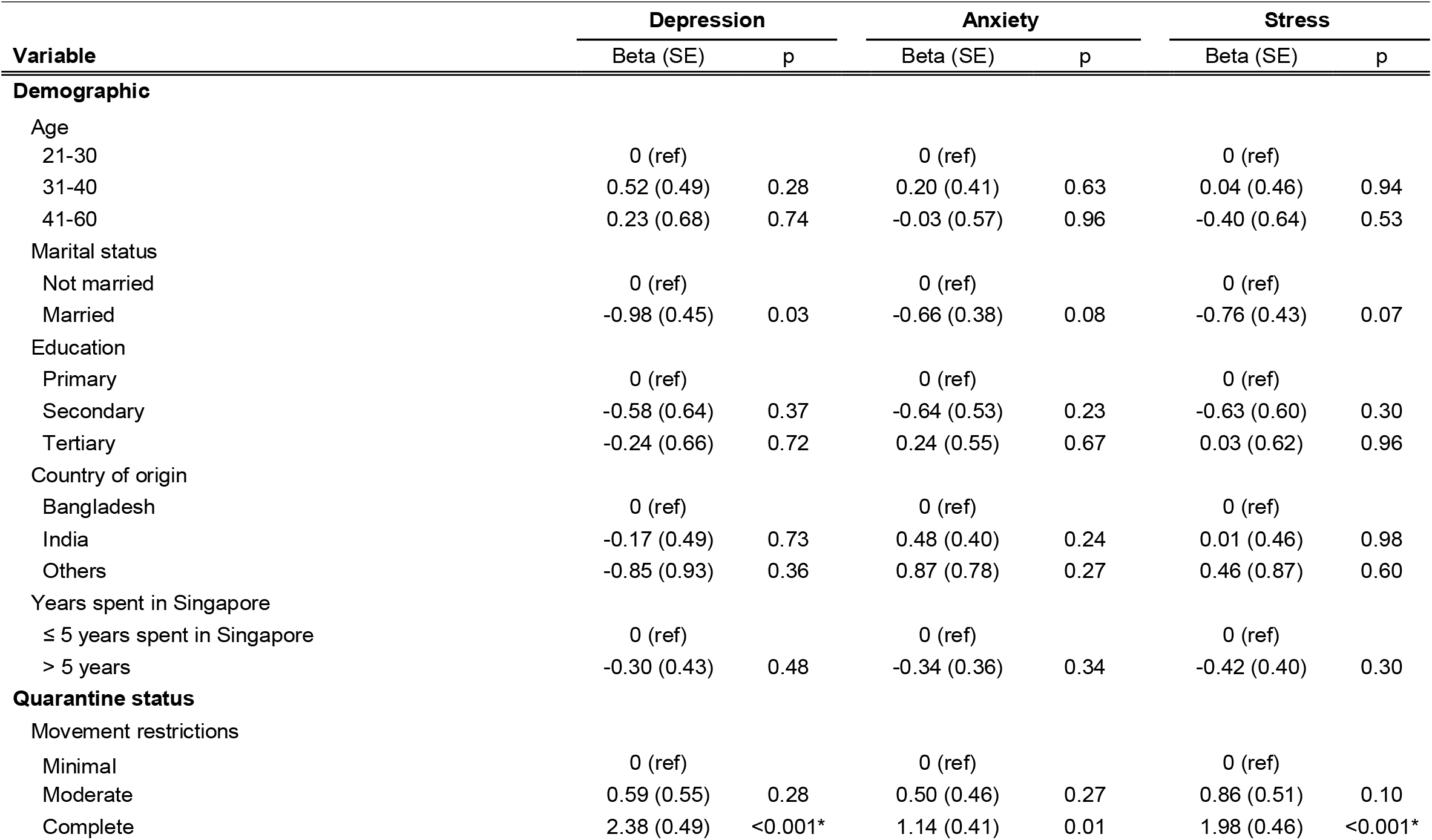
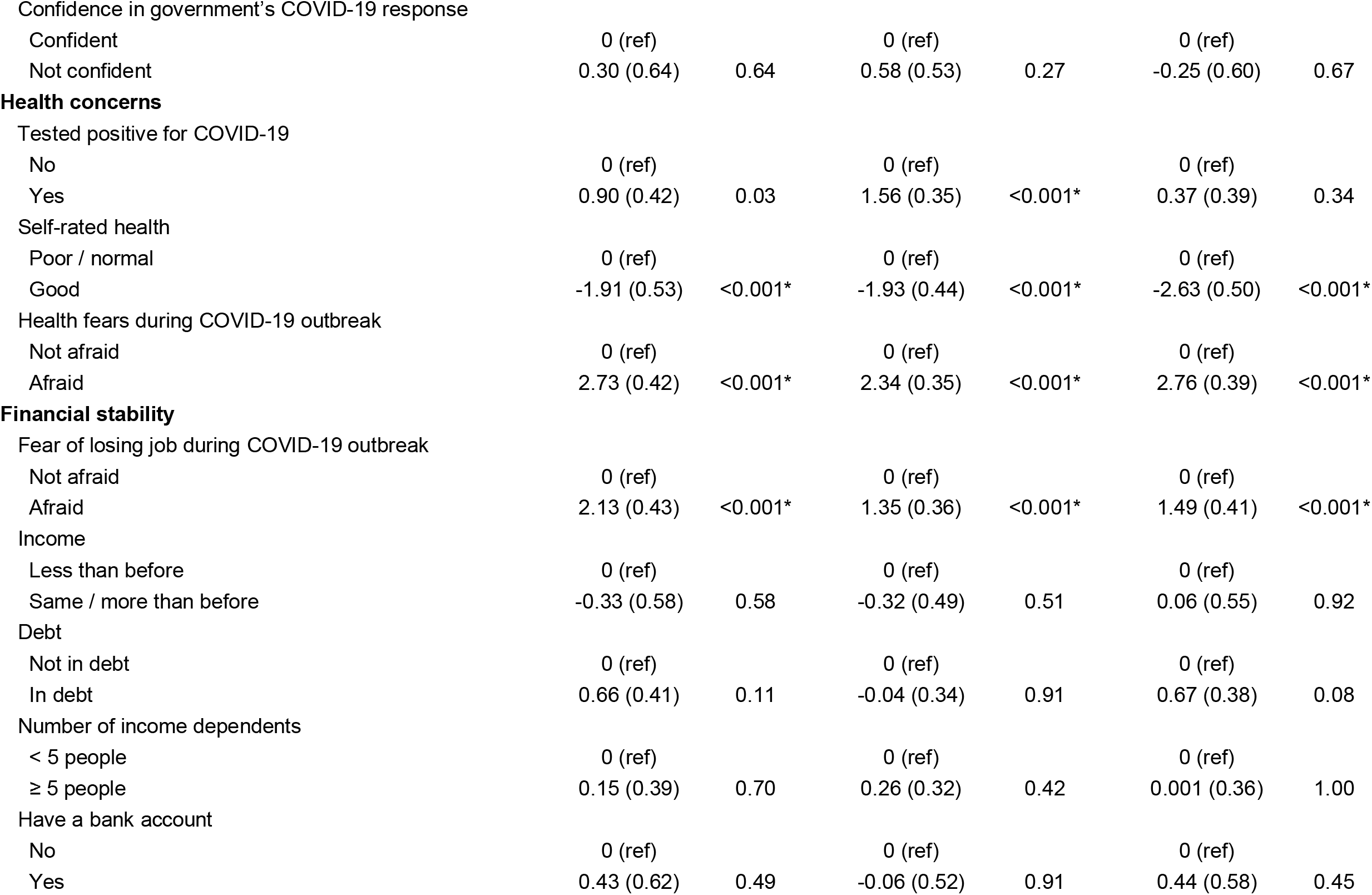
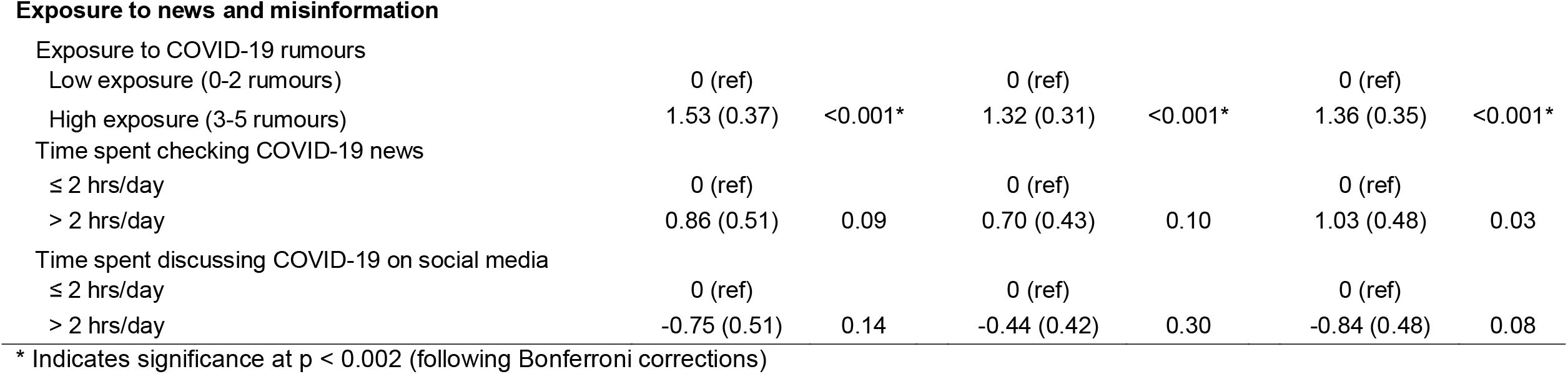
Linear regression models predicting depression, anxiety, and stress symptoms within a migrant worker community during the COVID-19 pandemic

| Variable | Depression |  | Anxiety |  | Stress |  |
| --- | --- | --- | --- | --- | --- | --- |
|  | Beta (SE) | p | Beta (SE) | p | Beta (SE) | p |
| **Demographic** |  |  |  |  |  |  |
| Age |  |  |  |  |  |  |
| 21-30 | 0 (ref) |  | 0 (ref) |  | 0 (ref) |  |
| 31-40 | 0.52 (0.49) | 0.28 | 0.20 (0.41) | 0.63 | 0.04 (0.46) | 0.94 |
| 41-60 | 0.23 (0.68) | 0.74 | -0.03 (0.57) | 0.96 | -0.40 (0.64) | 0.53 |
| Marital status |  |  |  |  |  |  |
| Not married | 0 (ref) |  | 0 (ref) |  | 0 (ref) |  |
| Married | -0.98 (0.45) | 0.03 | -0.66 (0.38) | 0.08 | -0.76 (0.43) | 0.07 |
| Education |  |  |  |  |  |  |
| Primary | 0 (ref) |  | 0 (ref) |  | 0 (ref) |  |
| Secondary | -0.58 (0.64) | 0.37 | -0.64 (0.53) | 0.23 | -0.63 (0.60) | 0.30 |
| Tertiary | -0.24 (0.66) | 0.72 | 0.24 (0.55) | 0.67 | 0.03 (0.62) | 0.96 |
| Country of origin |  |  |  |  |  |  |
| Bangladesh | 0 (ref) |  | 0 (ref) |  | 0 (ref) |  |
| India | -0.17 (0.49) | 0.73 | 0.48 (0.40) | 0.24 | 0.01 (0.46) | 0.98 |
| Others | -0.85 (0.93) | 0.36 | 0.87 (0.78) | 0.27 | 0.46 (0.87) | 0.60 |
| Years spent in Singapore |  |  |  |  |  |  |
| ≤ 5 years spent in Singapore | 0 (ref) |  | 0 (ref) |  | 0 (ref) |  |
| > 5 years | -0.30 (0.43) | 0.48 | -0.34 (0.36) | 0.34 | -0.42 (0.40) | 0.30 |
| **Quarantine status** |  |  |  |  |  |  |
| Movement restrictions |  |  |  |  |  |  |
| Minimal | 0 (ref) |  | 0 (ref) |  | 0 (ref) |  |
| Moderate | 0.59 (0.55) | 0.28 | 0.50 (0.46) | 0.27 | 0.86 (0.51) | 0.10 |
| Complete | 2.38 (0.49) | <0.001* | 1.14 (0.41) | 0.01 | 1.98 (0.46) | <0.001* |
| Confidence in government's COVID-19 response |  |  |  |  |  |  |
| Confident | 0 (ref) |  | 0 (ref) |  | 0 (ref) |  |
| Not confident | 0.30 (0.64) | 0.64 | 0.58 (0.53) | 0.27 | -0.25 (0.60) | 0.67 |
| **Health concerns** |  |  |  |  |  |  |
| Tested positive for COVID-19 |  |  |  |  |  |  |
| No | 0 (ref) |  | 0 (ref) |  | 0 (ref) |  |
| Yes | 0.90 (0.42) | 0.03 | 1.56 (0.35) | <0.001* | 0.37 (0.39) | 0.34 |
| Self-rated health |  |  |  |  |  |  |
| Poor / normal | 0 (ref) |  | 0 (ref) |  | 0 (ref) |  |
| Good | -1.91 (0.53) | <0.001* | -1.93 (0.44) | <0.001* | -2.63 (0.50) | <0.001* |
| Health fears during COVID-19 outbreak |  |  |  |  |  |  |
| Not afraid | 0 (ref) |  | 0 (ref) |  | 0 (ref) |  |
| Afraid | 2.73 (0.42) | <0.001* | 2.34 (0.35) | <0.001* | 2.76 (0.39) | <0.001* |
| **Financial stability** |  |  |  |  |  |  |
| Fear of losing job during COVID-19 outbreak |  |  |  |  |  |  |
| Not afraid | 0 (ref) |  | 0 (ref) |  | 0 (ref) |  |
| Afraid | 2.13 (0.43) | <0.001* | 1.35 (0.36) | <0.001* | 1.49 (0.41) | <0.001* |
| Income |  |  |  |  |  |  |
| Less than before | 0 (ref) |  | 0 (ref) |  | 0 (ref) |  |
| Same / more than before | -0.33 (0.58) | 0.58 | -0.32 (0.49) | 0.51 | 0.06 (0.55) | 0.92 |
| Debt |  |  |  |  |  |  |
| Not in debt | 0 (ref) |  | 0 (ref) |  | 0 (ref) |  |
| In debt | 0.66 (0.41) | 0.11 | -0.04 (0.34) | 0.91 | 0.67 (0.38) | 0.08 |
| Number of income dependents |  |  |  |  |  |  |
| < 5 people | 0 (ref) |  | 0 (ref) |  | 0 (ref) |  |
| ≥ 5 people | 0.15 (0.39) | 0.70 | 0.26 (0.32) | 0.42 | 0.001 (0.36) | 1.00 |
| Have a bank account |  |  |  |  |  |  |
| No | 0 (ref) |  | 0 (ref) |  | 0 (ref) |  |
| Yes | 0.43 (0.62) | 0.49 | -0.06 (0.52) | 0.91 | 0.44 (0.58) | 0.45 |
| **Exposure to news and misinformation** |  |  |  |  |  |  |
| Exposure to COVID-19 rumours |  |  |  |  |  |  |
| Low exposure (0-2 rumours) | 0 (ref) |  | 0 (ref) |  | 0 (ref) |  |
| High exposure (3-5 rumours) | 1.53 (0.37) | <0.001* | 1.32 (0.31) | <0.001* | 1.36 (0.35) | <0.001* |
| Time spent checking COVID-19 news |  |  |  |  |  |  |
| ≤ 2 hrs/day | 0 (ref) |  | 0 (ref) |  | 0 (ref) |  |
| > 2 hrs/day | 0.86 (0.51) | 0.09 | 0.70 (0.43) | 0.10 | 1.03 (0.48) | 0.03 |
| Time spent discussing COVID-19 on social media |  |  |  |  |  |  |
| ≤ 2 hrs/day | 0 (ref) |  | 0 (ref) |  | 0 (ref) |  |
| > 2 hrs/day | -0.75 (0.51) | 0.14 | -0.44 (0.42) | 0.30 | -0.84 (0.48) | 0.08 |

\* Indicates significance at $p < 0.002$ (following Bonferroni corrections)

**Figure 3.**
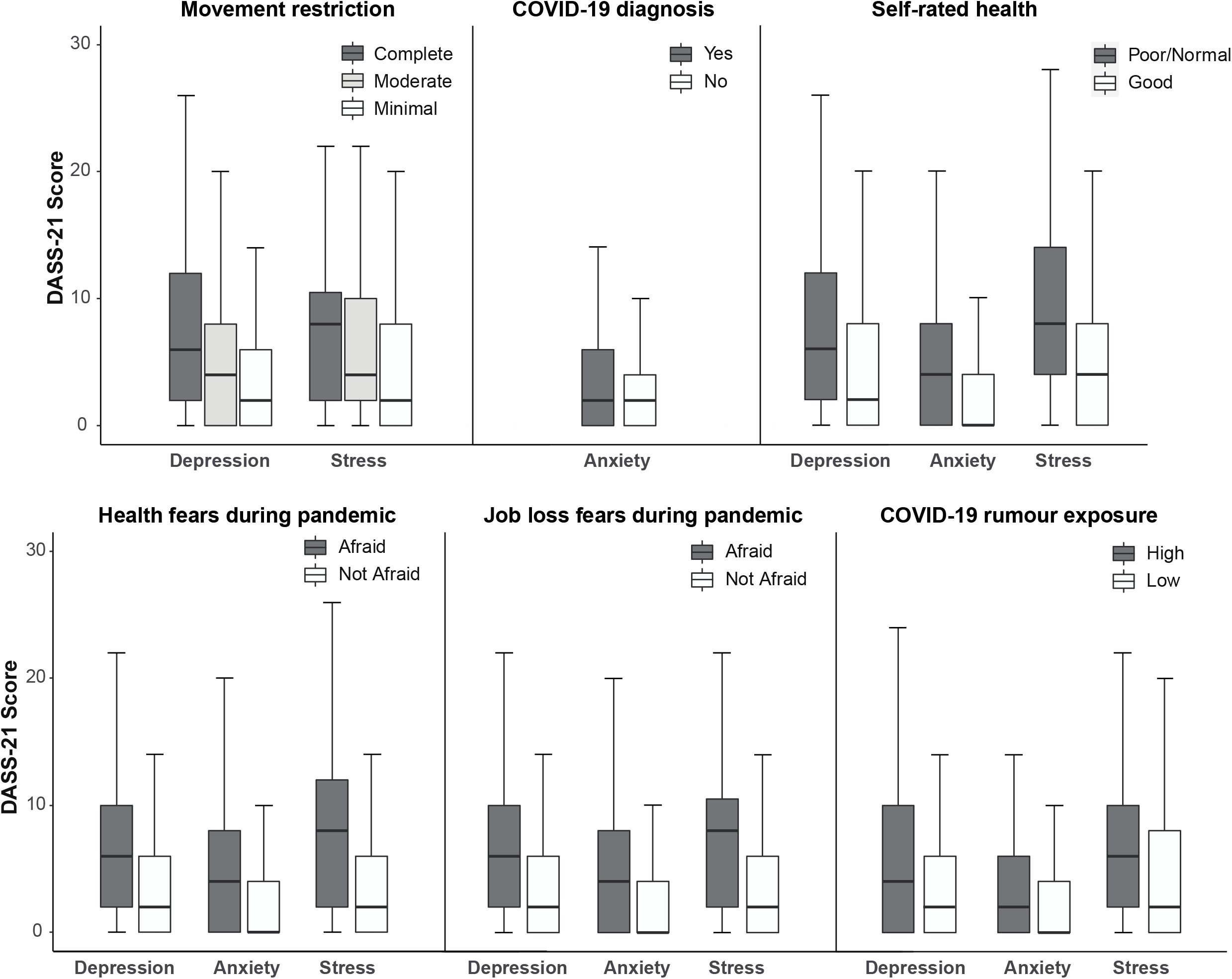
Box plots of participants’ scores on the Depression, Anxiety, and Stress Scale (DASS-21) as a function of: movement restrictions, testing positive for COVID-19, self-rated health, health fears during the pandemic, job loss fears during the pandemic, and COVID-19 rumour exposure.

In terms of health concerns, being diagnosed with COVID-19 predicted increased anxiety (b = 1.56, p < 0.001), but was not significantly associated with depression or stress levels. DASS-21 symptoms also tracked participants’ perception of the situation. Those who were fearful about their health during the pandemic reported more symptoms of depression, anxiety, and stress than those who were not fearful (depression: b = 2.73, p < 0.001; anxiety: b = 2.34, p < 0.001; stress: b = 2.76 p < 0.001). Conversely, participants who rated their health status as being good had fewer symptoms than those who rated their health as poor or normal (depression: b = −1.91, p < 0.001; anxiety: b = −1.93, p < 0.001; stress: b = - 2.63 p < 0.001). In terms of financial concerns, symptoms tracked participants’ subjective assessment of the situation, with higher depression, anxiety, and stress symptoms amongst participants who feared losing their job during the pandemic (depression: b = 2.13, p < 0.001; anxiety: b = 1.35, p < 0.001; stress: b = 1.49, p < 0.001). DASS-21 scores did not differ significantly as a function of income difference during COVID-19, having debt, the number of people dependent on participants’ incomes, or having a bank account in Singapore.

Higher DASS-21 symptoms were also observed amongst participants with higher exposure to COVID-19 rumours than those with lower exposure (depression: b = 1.53, p < 0.001; anxiety: b = 1.32, p < 0.001; stress: b = 1.36, p < 0.001). However, there was no significant relation between symptoms and the amount of time spent checking COVID-19 news or discussing COVID-19 on social media.

Finally, no sociodemographic factors significantly related to DASS-21 subscale scores.

## Discussion

Amidst growing concerns that the pandemic may broaden mental health inequalities,^20^ we conducted the first survey of depression, anxiety, and stress symptoms amongst low-waged migrant workers embroiled in large COVID-19 clusters. Through cross-sectional analyses, we identified several modifiable risk factors for poor mental health.

First, depression and stress symptoms differed as a function of government-imposed movement restrictions. On account of their high-density work and living environments (e.g., 25,000-person dormitory complexes), international migrant workers have come under severe forms of isolation protocols as governments acted to control large COVID-19 outbreaks.^13–15^ Against this backdrop, we recorded a higher number of mental health symptoms amongst participants undergoing complete restrictions – that is, prolonged in-dormitory or in-room quarantines to minimise cross-contact. In other words, while isolation of whole facilities may be expedient for limiting COVID-19 transmission (particularly when the pandemic started), this could come at the cost of increased mental health burden. Moving forward, other containment strategies could be prioritised: for example, implementing protocols that support the rapid detection and isolation of COVID-19 cases (e.g., frequent swab tests in migrant worker groups, or the use of digital contact tracing to track possible transmission), or reducing the density of migrant workers’ work and living environments.^14^

Aside from movement restrictions, anxiety symptoms were also elevated amongst participants who had tested positive for COVID-19. This echoes previous mental health surveys of COVID-19 patients,^11^ and is notable because several countries have reported higher prevalence rates of COVID-19 amongst migrant worker communities (relative to the general population in host countries).^9^

Depression, anxiety, and stress levels were further predicted by participants’ assessment of the situation, with symptoms elevated amongst those who: harboured fears about their health, perceived their health to be poor, or feared losing their jobs. Conversely, participants’ actual income, debt status, number of dependents, and access to banking facilities did not emerge as significant predictors. A possible explanation for these results is that depressed, anxious, and stressed individuals ruminate more, experiencing greater fear as a result. Alternatively, given that mental health symptoms tracked subjective self-reports more closely than objective markers (e.g., income), our results may reflect participants’ understanding of the situation, which in turn may be hampered by limited access to official COVID-19 messaging. This account is bolstered by our finding that participants with higher exposure to COVID-19 rumours reported elevated depression, anxiety, and stress symptoms. If supported by further studies, these results could shine a spotlight on the language and social barriers that many migrant workers encounter in their host countries, preventing access to official information sources that can provide assurance and counteract rumours.^4^ Follow-up research is needed to tease apart these accounts.

Although we have identified predictors of poor mental health, our documented rates of severe or extremely severe depression, anxiety, and stress levels are similar to those observed in the general population for the host country (Singapore).^19^ In other words, we found no evidence that migrant workers as a group had higher rates of mental health symptoms – despite their considerable vulnerability during the pandemic. This finding echoes previous studies describing the mortality advantage of international migrants outside the pandemic context,^21^ alluding to the resilience of the migrant worker community in the face of complex migration-related stressors. At the same time, we urge caution in interpreting these base rates. First, given restricted access to migrant workers amidst nation-wide lockdowns, we were unable to use a probabilistic sampling strategy. Consequently, our observed rates of symptomology may not represent prevalence rates for the migrant worker population as a whole. Second, while we did not observe elevated depression, anxiety, or stress levels within our sample, several newspaper articles have described multiple suicide attempts within the migrant worker dormitories.^22^ One way to reconcile these reports is by applying the diasthesis-stress model, which describes how mental disorders are the products of both situational circumstances and a person’s predispositions.^23^ Correspondingly, the possibility remains that although migrant workers showed resilience as a group, those who were pre-disposed to poor mental health may have been adversely affected by pandemic-related stressors, resulting in more suicide attempts. We urge further research studies to explore this possibility.

In presenting our findings, we note several study limitations. First, we captured participants’ mental health at only one time-point. Future studies should monitor participants’ depression, anxiety, and stress levels longitudinally, allowing us to assess the impact of changing movement restrictions over time. Second, the context for COVID-19 research is dynamic, and further studies are needed to assess whether our results generalize across stages of the pandemic and to other migrant groups. Finally, we used an epidemiological approach to identify risk factors for poor mental health. Although this method is commonly-used (particularly during the pandemic), the cross-sectional design precludes conclusions about causality.

Notwithstanding the study limitations, our study provides a rare window into the mental health of low-waged migrant workers during the COVID-19 pandemic. To the best of our knowledge, this is the first such survey since the pandemic started, and remains one of the largest mental health studies involving dormitory-housed migrant workers to date (even pre-pandemic). Although we observed resilience within our sample, the risk factors we identified underscore the need to ensure COVID-19 policies do not leave vulnerable groups behind.

## Data Availability

The data that support the findings of this study are available on request from the corresponding author (JCJL). The data are not publicly available due to IRB restrictions.

## Contributors

JCJL, PB and KD conceived of the study. YES, EYQT and JCJL carried out data collection and data analyses, accessed and verified the underlying data, and wrote the first draft of the manuscript. All authors contributed to editing and commenting on the final version.

## Declaration of interests

The authors declare no competing interests.

## Acknowledgements

This work was supported by a JY Pillay Global Asia Grant awarded to JCJL and KD (grant IG20-SG002). We are deeply grateful to the surveyors and translators who supported data collection; to hospital, government, and facility staff who granted us research access; to the HealthServe team for feedback during the design phase, and to Prof Jeannette Ickovics for comments on our draft. Above all, we are indebted to the migrant worker community for hosting us during a pandemic and for supporting this research.

## Notes

### Competing Interest Statement

The authors have declared no competing interest.

### Clinical Trial

NCT04448704

### Funding Statement

JY Pillay Global Asia Grant. The funders had no role in the design and conduct of the study; in data collection, analysis, and interpretation; or in manuscript preparation.

### Author Declarations

The study was approved by the Institutional Review Boards of the National University of Singapore and Singapore Health Services (2020/2559).

